# Effect of Dry Heat and Autoclave Decontamination Cycles on N95 FFRs

**DOI:** 10.1101/2020.05.29.20114199

**Authors:** Cole Meisenhelder, Loïc Anderegg, Andrew Preecha, Chiu Oan Ngooi, Lei Liao, Wang Xiao, Steven Chu, Yi Cui, John M. Doyle

## Abstract

Current shortages of Filtering Facepiece Respirators (FFRs) have created a demand for effective methods for N95 decontamination and reuse. Before implementing any reuse strategy it is important to determine what effects the proposed method has on the physical functioning of the FFR. Here we investigate the effects of two potential methods for decontamination; dry heat at 95 °C, and autoclave treatments. We test both fit and filtration efficiency for each method. For the dry heat treatment we consider the 3M 1860, 3M 1870, and 3M8210+ models. After five cycles of the dry heating method, all three FFR models pass both fit and filtration tests, showing no degradation. For the autoclave tests we consider the 3M 1870, and the 3M 8210+. We find significant degradation of the FFRs following the 121 °C autoclave cycles. The molded mask tested (3M 8210+) failed fit testing after just 1 cycle in the autoclave. The pleated (3M 1870) mask passed fit testing for 5 cycles, but failed filtration testing. The 95 °C dry heat cycle is scalable to over a thousand masks per day in a hospital setting, and is above the temperature which has been shown to achieve the requisite 3 log kill of SARS-CoV-2[1], making it a promising method for N95 decontamination and reuse.

## INTRODUCTION

The ongoing COVID pandemic has caused a shortfall of N95 respirators worldwide. There has been much work recently assessing various decontamination methods which may be employed in order to safely reuse these FFRs. A successful decontamination method should provide >3 log reduction of the virus and should not degrade either fit nor filtration of the FFR[2, 3]. Ideally, a decontamination method should also be easy to implement and have short cycle times. Few studies to date have investigated the effect of heat on SARS-CoV2 on the surface of FFRs. Recent work has shown that 70 °C for 60 minutes provides >3 log reduction for SARS-CoV2 in DMEM[1]. Similarly, one recent study has shown that an autoclave cycle of 15 minutes at 121 °C provides >4.6 log reduction of SARS-CoV2 on FFRs[4].

Previous studies of heat decontamination on FFR performance have been primarily performed in a temperature range of 70-75 °C, either below or very near the 70 °C for one hour limit [1, 5]. It should be noted that this limit was tested in a medium which may not accurately represent the real world local environment for the virus on FFRs, which would likely contain mucus or saliva, and which may change the temperature profile of inactivation. One study tested filtration efficiency of the 4C Air N95, showing no significant change at 100 °C for 30 min[6]. Autoclave studies have shown that molded (such as the 3M 1860 or 3M 8210+) models fail fit tests, while pleated models (such as the 3M 1870) pass fit tests[4, 7]. An additional study has shown that one pleated model (3M 1862+) passes filtration testing after multiple cycles of autoclave treatment[8, 9]. To date, no studies have shown passing of both fit and filtration tests for a single model.

Here we study the effects on fit and filtration for two methods of decontamination; dry heat at 95 °C and autoclave at 121 °C. We perform each heat treatment for five cycles, as previous studies have suggested that donning and doffing more than five times can reduce the fit factor for some mask models[10]. We find that the dry heat method satisfies both fit and filtration requirements for three models of FFR. Autoclave tests on two models reveal degradation either of the fit or filtration.

## HEATING PROTOCOL

The FFRs are placed into individual polypropylene food storage containers(ziplock medium squares) and heated in a convection oven (Despatch LAC1-38-8, 3.7 cu. Ft.). The individual containers are used as a method to prevent cross contamination between the FFRs. The process is similar to that described in our previous publication[11], omitting the moisture source to produce dry heat. For temperature validation, each container’s lid is modified to contain a temperature and humidity sensor(SEK-SHT31-Sensors logged with SEK-SensorBridge). Figure 1 shows the temperature as a function of time inside the containers. Each cycle lasts 40 min, due to a 10 minute warm-up time. After 40 minutes, the FFRs are taken out of the oven and the lids are opened to expedite the cooling. After each heating cycle, both a visual inspection and a quantitative fit test of the masks is performed.

**Figure. 1.**
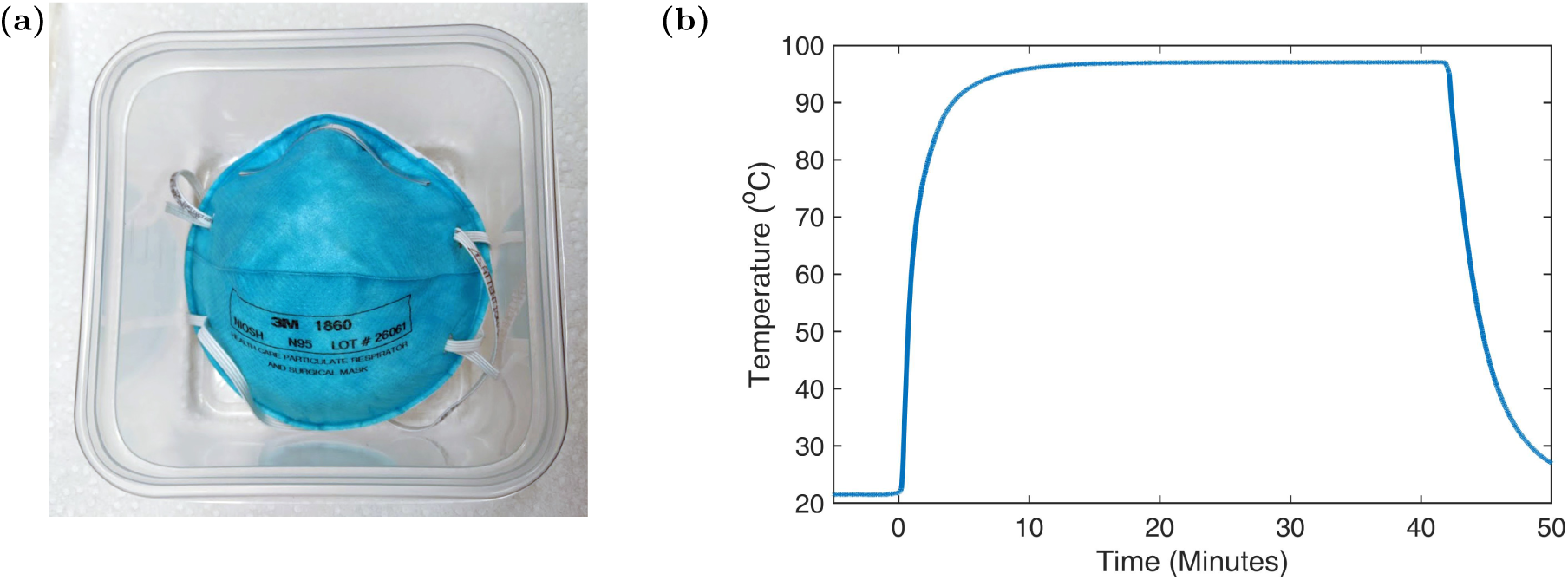
(a) Each N95 FFR was placed into a plastic container to prevent cross contamination. (b) Temperature profile of the mask during the heating cycle. The masks reach within two degree Celsius of the setpoint within ~ 10 minutes.

## AUTOCLAVE PROTOCOL

Testing of three FFR models with the autoclave is performed as follows. Three models (3M 8210, 8210+, and 1870) are placed into lidless polypropylene containers. Autoclave tape is placed on the bottom and sides of the polypropylene boxes as a visual indication that a temperature of 121 °C is achieved during the autoclave cycle. The boxes and FFR are placed in a Nalgene tray and positioned roughly in the center of the chamber of a Getinge 533LS steam sterilizer. The door of the autoclave is then closed and the Vac20/20 cycle was selected. This cycle consists of a 10 min prep period (where steam is generated and air pumped out), a 20 min exposure at 121 °C, and a 20 min vacuum drying period. After waiting a few minutes after the drying step, the door is opened, taking care that the operator is not directly in front of the door so as to not be exposed to residual steam and heat. The tray is removed and allowed to cool down to room temperature.

Masks are qualitatively assessed for overall look and structure. Firstly, the elastic straps are tested for elasticity and brittleness. This is done by making a 1 mm diameter mark on a rubber band strap using a permanent marker. Before and after each cycle, the bands are stretched until resistance is met. The stretched length of the mark in these states is then measured. Additionally, the straps are stretched over the tester’s head 10 times as if the masks were to be donned and doffed, with the texture and feel of the rubber noted after each extension. The foam around the nose of the FFR is assessed for flexibility and brittleness. This is done by visual inspection of the foam cell size as well as tactile examination of softness. Special care is taken in determining if the foam padding is delaminating after each cycle. Lastly, a quantitative fit test is performed with the results shown in figure 2. Once the masks are fit checked, they are placed back into the autoclave as described and the sterilization is repeated for a total of 5 cycles.

**Figure. 2.**
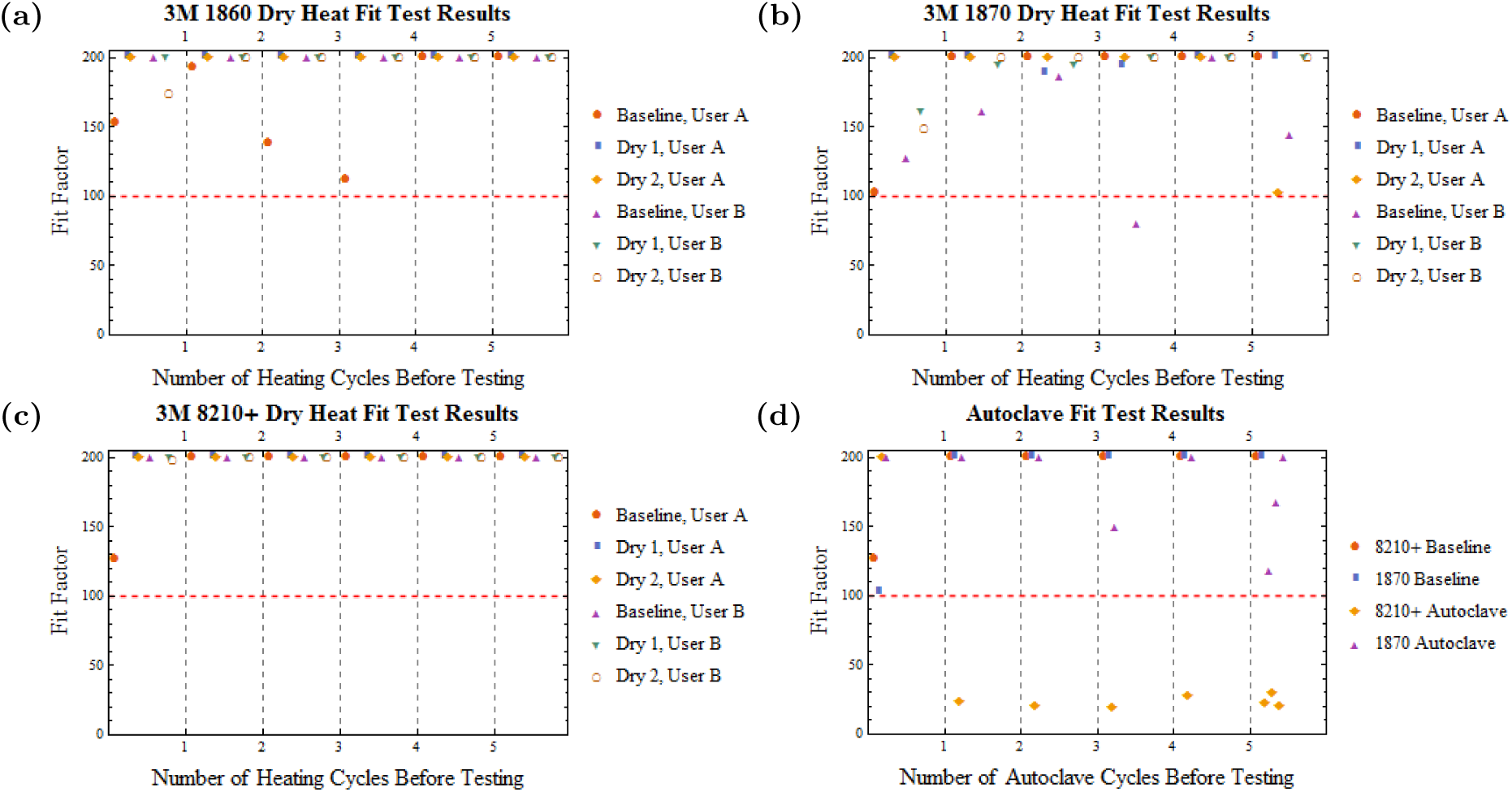
Quantitative fit test results between each dry heat cycle for the 3M (a) 1860, (b) 1870, and (c) 8210+. (d) Fit test results following the autoclave cycles.

## FIT TESTING METHODS

Fit testing was performed with a TSI PortaCount Respirator Fit Tester 8038 using NaCl particles produced by a particle generator (TSI 8026) and the OSHA modified ambient aerosol Condensation Nuclei Counter (CNC) Quantitative Fit Test for Filtering Facepiece Respirators. The test is composed of four components; bending at the waist for 50 seconds, talking for 30 seconds, turning the head side to side for 30 seconds, and lifting the head up and down for 39 seconds. Each segment produces a quantitative fit factor and an overall fit factor is computed as the geometric mean of these scores. A passing score is awarded for an overall fit factor ≥100, even if an individual category is below 100. The range of the system is limited to a maximum score of 200+. For each measurement the user donned their mask and performed a qualitative user seal check to identify any noticeable leaks and adjust for any leaks. After the fit tests the user doffed the mask to simulate all stages of mask usage.

Fit tests for the dry heat method were performed on two users, designated throughout as User A, and User B. Each user performed fit tests with three FFR models, the 3M 1860 surgical N95, the 3M 8210+, and the 3M 1870 surgical N95. For all models each user performed six fit tests with a control mask to provide a baseline for comparison and to simulate an initial test followed by another test after each of five treatment cycles. User A performed baseline fit tests with the same mask models under the same conditions for a prior study[11], and so this data was reused to conserve masks. For the test group, each user performed fit tests on two masks of each model before beginning the heat treatments and after each heating cycle, for five cycles. For the autoclave treatment only User A performed fit tests for the two models tested (3M 8210+ and 3M 1870). For each mask model the user performed fit tests with a single mask in the test group before the autoclave treatment began and between each treatment cycle. Due to the low mask number, the masks were fit tested three times after the final autoclave cycle to capture any variance in the users ability to form a seal. The number of masks used in each treatment method were chosen to conserve masks during the current shortage.

## FILTRATION TESTING METHODS

The filtration testing protocol was similar to that previously described in (Anderegg 2020[11]). In brief, a TSI Inc. Automated Filter Tester 8130A was used with 0.26 μm (mass mean diameter) NaCl particles as the aerosol source at a flow rate of 85 L/min. This system is able to determine both the filtration efficiency as well as the pressure drop across the FFR. We report two filtration numbers, both the initial filtration as well as the filtration after a 60 minute loading cycle. This loading cycle is used to mimic the effects of soiling of the mask as the mask is used. Per NIOSH standards, the N95 rating requires this filtration efficiency remains above 95% during the entire loading cycle.

## RESULTS AND DISCUSSION

The filtration results are shown in Table 1. The filtration of each mask type is tested without any treatment as a baseline value for that mask type. After 5 decontamination cycles (either dry heat or autoclave) the filtration is tested. Each measurement is an average of two masks. We find the mask to mask filtration efficiency to be extremely constant. For the dry heat, we find that after 5 cycles, all three masks remain well above the 95% filtration standard. Both the 3M 1870 and 8210+ showed essentially no degradation in both initial filtration and minimum filtration during loading. The 1860 mask shows a small amount of filtration degradation during the loading test, but remained well above the 95% standard. However, we see significant degradation of both the pleated and molded models after the autoclave cycles. The 1870 fell below the 95% filtration requirement and the 8210, while technically above the minimum filtration limit, nearly failed. The pressure drop of all masks remained far below the maximum resistance to airflow specified by NIOSH ([19] 42 CFR 84.180).

**Table. I.**
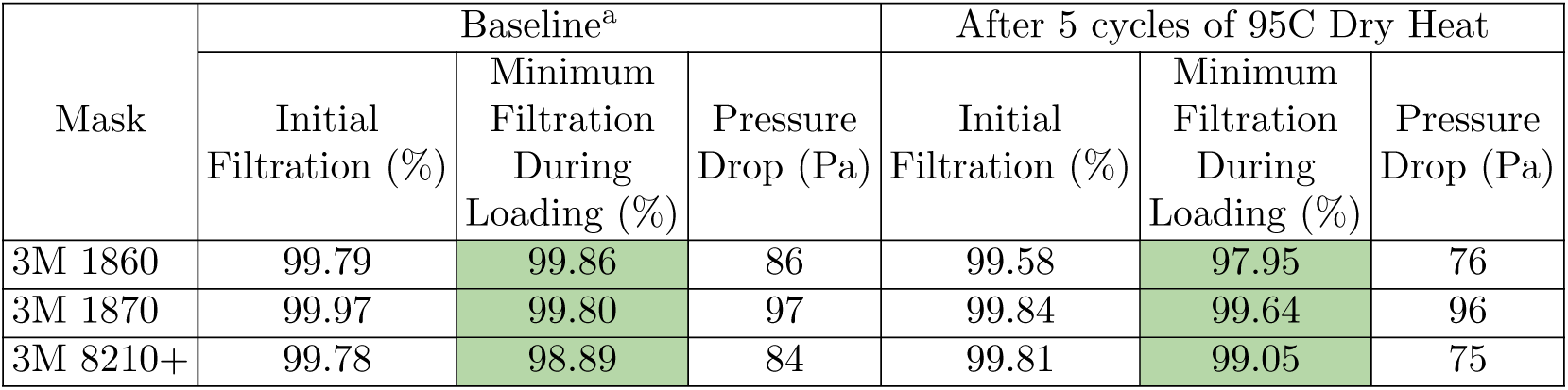
Filtration testing results for dry heat cycles

**Table. II.** Filtration testing results for autoclave cycles

| Mask | Baseline |  |  | After 5 Autoclave Cycle |  |  |
| --- | --- | --- | --- | --- | --- | --- |
|  | Initial Filtration (%) | Minimum Filtration During Loading (%) | Pressure Drop (Pa) | Initial Filtration (%) | Minimum Filtration During Loading (%) | Pressure Drop (Pa) |
| 3M 1870 | 99.97 | 99.80 | 97 | 98.28 | 93.89 | 95 |
| 3M 8210+ | 99.78 | 98.89 | 84 | 97.8 | 95.04 | 74.5 |

## FIT TEST RESULTS

The results of the fit tests for the dry heat treatments are shown in Figure 2(a-c), broken up for each FFR model. Each model passed fit all fit tests after heat treatment without significant degradation. It should be noted that User B failed the fit test for one of the baseline measurements with the 3M 1870 model, indicating that this model is not ideally suited to their face, however they saw no failures in the test group. The fit test results for the autoclave tests are summarized in Fig 2(d). As demonstrated in prior studies[4, 7, 8], the molded 8210+ model failed fit tests after a single cycle. The pleated 1870 model passed all fit tests, while only showing a slight decrease in fit factor after all five cycles, which was not significant enough to cause failure.

## QUALITATIVE ASSESSMENTS

After dry heating, there was noticeable blurring of the manufacturer’s ink in all of the FFR models after a single cycle, similar to that observed for the autoclaved masks and in other studies[11]. Additionally, the 3M 1870 sees the same delamination of the foam nose piece that is observed with the autoclave treatment and previous heat studies[11], however this does not interfere with the fit tests.

For all FFR models tested there is no degradation of the elasticity of the straps after each autoclave cycle. This is tested by measuring the extension factor of the straps. Additionally, each of the straps withstood 50 donning and doffing motions without degradation. There is no brittleness or loosening of the straps observed, and the stitching and staples attaching the straps are unaffected after 5 autoclave cycles.

Blurring of the manufacturer’s ink for each model is also noted after a single autoclave cycle, but no outward signs of damage or degradation are noted to the materials of the masks after 5 autoclave cycles. Slight delamination of the foam nosepiece as noted after one autoclave cycle with the 1870 masks, but this delamination does not increase after subsequent cycles. The molded masks (8210 and 8210+) experience some shrinkage after 5 cycles. The full extent of this shrinkage is not characterized in this report. The pleated 1870 experience no visible shrinkage.

## CONCLUSION

We have shown that for three common 3M FFRs (1860, 1870, 8210+) 95 °C dry heat can be applied for 30 minutes for at least 5 cycles without significant degradation of either fit or filtration. This is a scalable and simple heating option that may be implemented for decontamination of FFR in a wide range of healthcare settings. We find however that the masks were not able to survive autoclave cycles. This is contrary to some recent studies. One possible reason for this discrepancy is differences in filtration testing methodology.

## Data Availability

All data associated with this work is included in the manuscript.

## ACKNOWLEDGMENTS

We acknowledge and appreciate the support of the Heising-Simons foundation.

## Footnotes

1 These baseline vales were previously measured and reported by the authors in [11]

